# Brain-derived Tau for Monitoring Brain Injury in Acute Ischemic Stroke

**DOI:** 10.1101/2023.11.18.23298728

**Authors:** Naomi Vlegels, Fernando Gonzalez-Ortiz, Nicoló Luca Knuth, Nada Khalifeh, Benno Gesierich, Franziska Müller, Philipp Müller, Matthias Klein, Konstantinos Dimitriadis, Nicolai Franzmeier, Thomas Liebig, Marco Duering, Paul Reidler, Martin Dichgans, Thomas K. Karikari, Kaj Blennow, Steffen Tiedt

## Abstract

The evolution of infarcts varies widely among patients with acute ischemic stroke (IS) and influences treatment decisions. Neuroimaging is not applicable for frequent monitoring and there is no blood-based biomarker to track ongoing brain injury in acute IS. Here, we examined the utility of plasma brain-derived tau (BD-tau) as a biomarker for brain injury in acute IS. We conducted the prospective, observational Precision Medicine in Stroke [PROMISE] study with serial blood sampling upon hospital admission and at days 2, 3, and 7 in patients with acute ischemic stroke (IS) and for comparison, in patients with stroke mimics (SM). We determined the temporal course of plasma BD-tau, its relation to infarct size and admission imaging-based metrics of brain injury, and its value to predict functional outcome. Upon admission (median time-from-onset, 4.4h), BD-tau levels in IS patients correlated with ASPECTS (*ρ*=-0.21, *P*<.0001) and were predictive of final infarct volume (*ρ*=0.26, *P*<.0001). In contrast to SM patients, BD-tau levels in IS patients increased from admission (median, 2.9 pg/ml [IQR, 1.8-4.8]) to day 2 (median time-from-onset, 22.7h; median BD-tau, 5.0 pg/ml [IQR, 2.6-10.3]; *P*<.0001). The rate of change of BD-tau from admission to day 2 was significantly associated with collateral supply (*R^2^*=0.10, *P*<.0001) and infarct progression (*ρ*=0.58, *P*<.0001). At day 2, BD-tau was predictive of final infarct volume (*ρ*=0.59, *P*<.0001) and showed superior value for predicting the 90-day mRS score compared with final infarct volume. In conclusion, in 502 patients with acute IS, plasma BD-tau was associated with imaging-based metrics of brain injury upon admission, increased within the first 24 hours in correlation with infarct progression, and at 24 hours was superior to final infarct volume in predicting 90-day functional outcome. Further research is needed to determine whether BD-tau assessments can inform decision-making in stroke care.

## INTRODUCTION

Ischemic stroke remains a leading cause of death and long-term disability worldwide,^1^ despite major advancements in reperfusion therapies.^2,3^ While neuroimaging modalities have expanded patient eligibility for reperfusion therapies by estimating ischemic core and salvageable tissue,^4–7^ their assessments are mostly single-timed. Currently available clinical algorithms lack the capacity to continuously track the dynamic evolution of how the primary core progresses to a final infarct, which, however, is a major determinant of functional outcome.^8–10^ Monitoring infarct trajectories could support therapeutic decision-making in patients with large-vessel occlusion stroke and unveil determinants of stroke progression, aiding in patient selection for trials evaluating cytoprotection^11^ and targeting clinically ineffective reperfusion.^12^ Contrary to neuroimaging, a blood-based biomarker would allow to monitor infarct growth with high frequency: in clinical routine, blood tests already support the monitoring of injuries of most organs, except of the brain. Such a molecular biomarker might also provide valuable biological insights including on the actual timing and extent of neuronal death.

Previously studied blood-based biomarkers such as Neurofilament Light Chain (NfL),^13^ neuron-specific enolase (NSE),^14^ glial fibrillary acidic protein (GFAP),^15,16^ and S 100 calcium-binding protein B (S100B)^17^ either failed to capture the extent of brain injury within the acute phase of stroke or lack specificity. Brain-derived tau (BD-tau) is a novel blood-based biomarker that quantifies tau protein originating specifically from the central nervous system (CNS).^18^ Unlike NfL, which serves as a core structural protein within axons,^19^ Tau dynamically binds to microtubules, mainly in dendrites and distal axons,^20,21^ and may thus be released into the extracellular space more quickly upon neuronal death.^22,23^ Indeed, in recent studies on neurodegenerative diseases^18^ and traumatic brain injury^24^, BD-tau outperformed established biomarkers of brain injury including NfL in the early detection of CNS pathology.

Here, we set out to determine whether BD-tau is rapidly released after stroke onset, correlates with the extent of brain injury in the acute phase of ischemic stroke, and is predictive of long-term functional outcome.

## METHODS

### Study design, patients, and procedures

The Precision Medicine in Stroke (PROMISE) study was a prospective observational study (ClinicalTrials.gov number: NCT05815836) with the aim to identify novel blood-based biomarkers for stroke. Patients were recruited in the emergency department of the LMU University Hospital, a comprehensive stroke center in Munich, Germany, between October 2013 and February 2021. Further details on screening of patients, procedures intended to ensure a representative sample of eligible patients, and criteria for inclusion and exclusion are provided in the eMethods in the Supplement. Patients were eligible if they presented with rapidly developing clinical signs suggestive of stroke within 24 hours of symptom onset and were at least 18 years of age. Patients were excluded if they had a stroke, myocardial infarction, other vascular event, or major surgery in the four weeks prior to admission. Among initially eligible patients with ischemic stroke or stroke mimic, we selected those who had blood samples collected upon admission (day 1), the next morning (day 2), day 3, and day 7. Details regarding the collection and management of data including of functional outcome at follow-up, using the modified Rankin Scale (mRS) score, are provided in the eMethods in the Supplement. Healthy controls were recruited through a single outpatient clinic at LMU University Hospital (eMethods in the Supplement). All patients as well as healthy controls gave informed consent in accord with local ethical approvals (Ref. No: 121-09 and 23-0143). Details regarding deferred consent are provided in eMethods in the Supplement. The study was conducted in accordance with the Declaration of Helsinki and is reported according to the Standards for Reporting Observational Studies (STROBE) guidelines.

### Plasma processing and BD-tau assay

Details on blood sampling and processing are provided in the eMethods in the Supplement. Plasma BD-tau was quantified on the Simoa HD-X platform (Quanterix) at the University of Gothenburg, Mölndal, Sweden, applying a previously described protocol^18^ (eMethods in the Supplement). Plasma samples were diluted four times with the Homebrew buffer (101556, Quanterix, MA, USA) before measurement. Stated concentration values were adjusted for the pre-measurement dilution. Intra- and inter-run repeatability was calculated using two internal quality controls: variation in the whole cohort was <10 % (eTable 1 and eFigure 1 in the Supplement). Experimenters were blinded to all clinical data.

### Neuroimaging

Multimodal CT, obtained as part of clinical routine upon hospital admission, followed a standard protocol for non-contrast CT, CT angiography, and CT perfusion and was leveraged to assess the Alberta Stroke Program Early CT Score (ASPECTS)^25^ including for the posterior circulation,^26^ collateral supply according to Tan et al.^27^ and Menon et al.,^28^ and ischemic core volume (cerebral blood volume <1.2 mL/100 mL).^29^ Final infarct volume was quantified using delayed diagnostic scans (at least 48 hours after stroke onset, mean time from onset to imaging: 4 days), either CT or MRI. Further details on the assessment of imaging-based metrics of brain injury are provided in the eMethods in the Supplement.

### Statistical analyses

Comparisons between ischemic stroke, stroke mimics and healthy controls were tested with linear models, adjusted for age and sex and post-hoc Tukey HSD tests (corrected for multiple testing). Linear mixed models were used (‘lme4’ package in R)^30^ to test the temporal course of BD-tau in ischemic stroke. Standardized beta and 95% confidence intervals (CI) are reported. The difference between relative changes of BD-tau and NfL levels in percentage in the acute phase were tested with a paired-sample t-test. The change in BD-tau levels from admission to day 2 was calculated and corrected for the hours between sampling on admission and day 2 for each individual patient and termed ΔBD-tau. To determine the relation between BD-tau and imaging-based metrics of brain injury, we used spearman correlation for continuous variables and analysis for variance (ANOVA) for categorical variables. To test whether BD-tau levels were different with the occurrence of either secondary intracerebral hemorrhage, recurrent ischemic stroke, or hemorrhagic transformation, we used an ANOVA, adjusted for infarct volume. The relationship between BD-tau and the National Institutes of Health Stroke Scale (NIHSS) score was determined using spearman correlation. To determine the relationship of BD-tau with the mRS score at 7 and 90 days after stroke (as dependent variable) we used ordinal (for the mRS score distribution) and binomial (for the rate of functional independence defined as an mRS score of 0-2) univariable and multivariable logistic regression adjusting for potential baseline confounder variables (age, sex, hypertension, pre-stroke mRS). Odds ratios (OR) and 95% CI are reported. A *P* value <.05 was considered statistically significant and all tests were performed two-sided. Random forest regression was used to assess conditional variable importance in multivariable models for the dependent variables BD-tau (day 1 and ΔBD-tau) and 90-day functional outcome. Details of the statistical methods are provided in the eMethods in the Supplement.

## RESULTS

### Study population

The enrollment and eligibility of patients in the PROMISE study are described in eFigure 2: 502 patients with ischemic stroke, 51 patients with stroke mimics (57% epileptic seizures, eTable 2 in the Supplement), and 102 healthy controls were included for analysis. Among patients with ischemic stroke, BD-tau levels were available for all 502 patients at admission and day 2 (100%), for 498 patients at day 3 (99.2%), 500 patients at day 7 (99.6%), and 124 patients at day 90 (24.7%). Data on functional outcome were available for 497 patients at day 7 (99.0%) and 363 patients at day 90 (72.3%). Final infarct volume data were available on 487 patients (97.0%).

The median age of patients with ischemic stroke was 76 years and 208 (41%) of them were women. The median time from onset to admission was 4.4 hours (interquartile range [IQR], 2.1 to 7.7). The median time from onset to sample collection at day 2 and 3 was 22.7 hours (IQR, 19.5 to 25.3) and 46.6 hours (IQR, 43.4 to 49.1), respectively. The median NIHSS score upon admission was 6 (IQR, 3 to 13) and the median final infarct volume was 9.4 ml (IQR, 1.9 to 36.7) (Table). The sample was representative of patients presenting with acute ischemic stroke with regards to age and stroke severity while showing higher than average rates of reperfusion treatments (eTable 3 in the Supplement).

**Table:** Characteristics of healthy controls and patients at baseline.

**Table: Characteristics of healthy controls and patients at baseline.**
| Characteristics | IS<br>N = 502 | SM<br>N = 51 | HC<br>N = 102 | *P* value |
| --- | --- | --- | --- | --- |
| Age, median (IQR) [years] | 76 (66, 83) | 78 (64, 85) | 75 (71, 78) | .66 |
| Female, No. (%) | 208 (41) | 24 (47) | 45 (44) | .68 |
| Medical history, No. (%) |  |  |  |  |
| Hypertension | 390 (78) | 37 (73) | 77 (75) | .66 |
| Diabetes mellitus | 107 (21) | 10 (20) | 10 (9.8) | .03 |
| Dyslipidemia | 12 (2.4) | 0 (0) | 1 (1.1) | .67 |
| Atrial fibrillation | 124 (25) | 12 (24) | 7 (6.9) | <.001 |
| Symptom onset to first sampling, median (IQR) [min] | 263 (125 – 460) | 151 (95 – 510) | NA | .47 |
| Pre-stroke mRS score, median (IQR) | 0 (0 – 1) | NA | NA | NA |
| Baseline NIHSS score, median (IQR) | 6 (3 – 13) | NA | NA | NA |
| ASPECTS, No. (%) |  |  |  |  |
| 0-2 | 3 (0.6) | NA | NA | NA |
| 3-5 | 16 (3.2) | NA | NA | NA |
| 6-8 | 76 (15) | NA | NA | NA |
| 9-10 | 364 (73) | NA | NA | NA |
| Intravenous alteplase treatment, No. (%) | 244 (48.6) | NA | NA | NA |
| Endovascular treatment, No (%) | 175 (34.8) | NA | NA | NA |
| Successful recanalization, No (%) | 150 (85.7) | NA | NA | NA |
| Infarct volume, median (IQR) | 9.35 (1.9 – 36.7) | NA | NA | NA |

IS, ischemic stroke; SM, Stroke mimics; HC, healthy controls; mRS, modified Rankin Scale; IQR,
interquartile range; NIHSS, National Institutes of Health Stroke Scale; ASPECTS, Alberta Stroke Program
Early CT Score; min, minutes; NA, not applicable.

### Temporal course of BD-tau after ischemic stroke

In patients with acute ischemic stroke, BD-tau levels increased from admission (median, 2.9 pg/ml [IQR, 1.8 to 4.8]) to day 2 (median, 5.0 pg/ml [IQR, 2.6 to 10.3]; *P*<.0001) (Figure 1A) and continued to rise at day 3 (median, 6.1 pg/ml [IQR, 3.2 to 15.0]) and day 7 (median, 7.2 pg/ml [IQR, 3.4 to 20.2]) followed by a decrease at day 90 (median, 2.2 pg/ml [IQR, 1.5 to 3.0]) (between any two time points: *P*<.0001). Patients with ischemic stroke showed higher BD-tau levels upon admission, at day 2, day 3, and day 7 compared with healthy controls (median, 1.8 pg/ml [IQR, 1.3 to 1.9]; all *P*<.0001), also when adjusting for age and sex (all *P*<.0001). Compared to patients with stroke mimics, BD-tau levels in patients with ischemic stroke were significantly higher at day 2, day 3, and day 7 (all *P*<.05) (Figure 1A), also when adjusting for age and sex (all *P*<.05).

**Figure 1.**
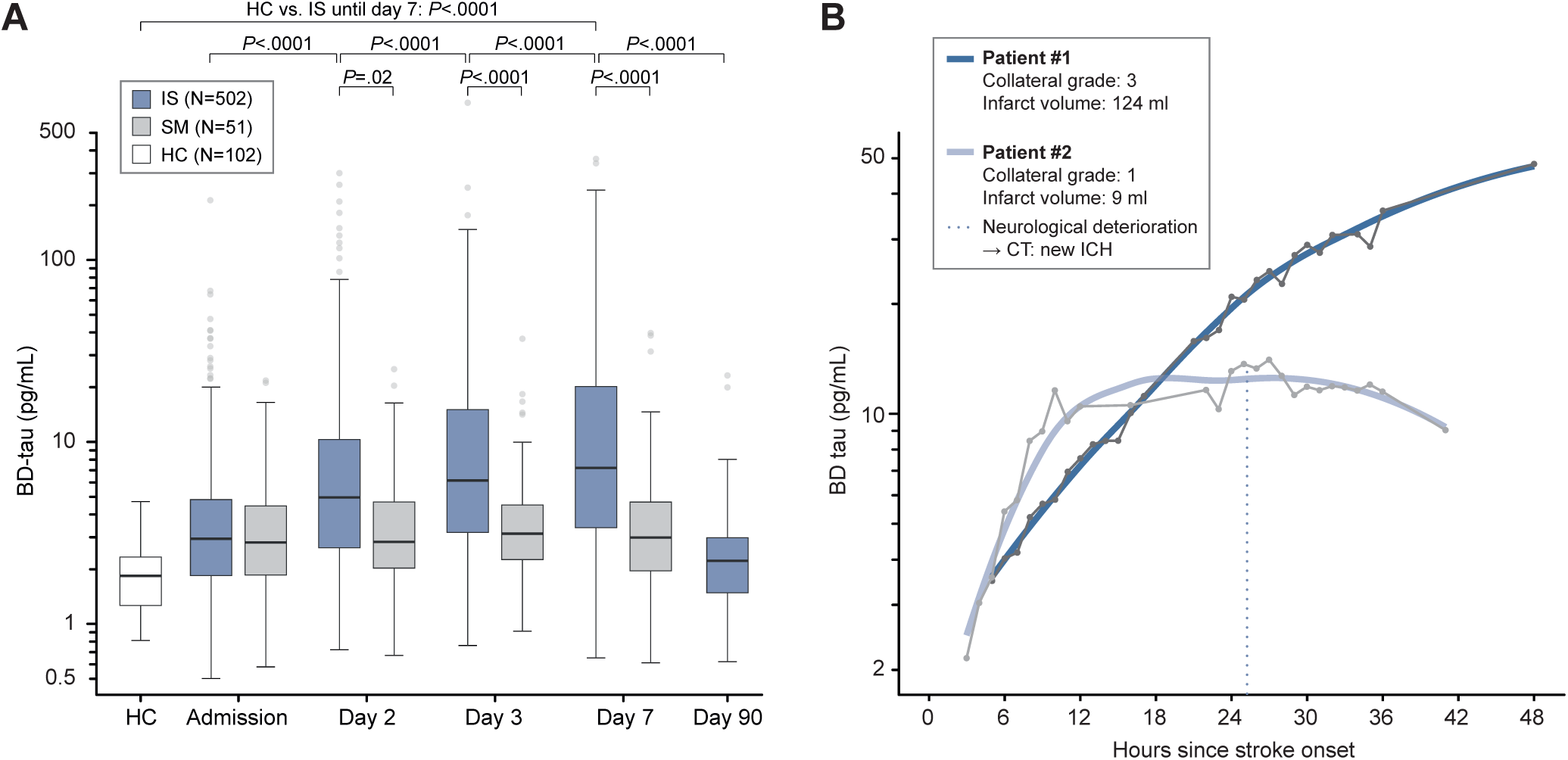
Temporal course of plasma BD-tau levels after stroke onset. (**A**) Plasma levels of BD-tau in patients with ischemic stroke over time in comparison to patients with stroke mimics and healthy controls. The boxes represent (from bottom to top) the first quartile, the median and the third quartile. The upper whiskers indicate the third quartile plus 1.5 times the interquartile range while the lower whiskers indicate the first quartile minus 1.5 times the interquartile range. *P* values were calculated using a linear mixed model (to compare time points within the stroke cohort) and analysis of variance with post-hoc Tukey HSD tests (to compare patients with ischemic stroke with healthy controls and stroke mimics). (**B**) Shown are serial high-frequency assessments of BD-tau levels of two patients in the first 48 hours after stroke onset. Plasma samples were collected hourly, except between 3AM and 5AM in the morning and during diagnostic procedures. BD-tau levels are indicated by dots, the blue lines indicate the fit derived from unbiased local polynomial regression. BD-tau, brain-derived tau; IS, ischemic stroke; SM, stroke mimics; HC, healthy controls; CT, computerized tomography; ICH, intracerebral hemorrhage.

To determine whether BD-tau levels start to rise early after stroke onset and to capture the real-time evolution of BD-tau in the context of collateral supply and final infarct volume, we performed hourly blood sampling in two patients from admission until 36 hours (eMethods in the Supplement): a rise in BD-tau levels was observed from the first assessments three and five hours after onset. Patient #1, demonstrating high collateral supply upon admission, showed continuously increasing BD-tau levels until at least 48 hours after onset. In contrast, patient #2, demonstrating low collateral supply, displayed a more rapid hourly change of BD-tau levels compared to patient #1 and reached a plateau already ten hours after onset. This was followed by another rise, which preceded the diagnosis of secondary intracerebral hemorrhage. During the first 36 hours after onset, peak BD-tau levels were considerably higher in patient #1 (final infarct volume: 124 ml) compared with patient #2 (final infarct volume: 9 ml) (Figure 1B).

### Relation of BD-tau to imaging-based metrics of infarct progression and final infarct size

Upon admission, BD-tau levels were significantly correlated with non-contrast CT-based ASPECTS (*ρ*=-0.21, *P*<.0001) and predictive of final infarct volume as assessed by delayed neuroimaging (*ρ*=0.26, *P*<.0001) (eFigure 3 in the Supplement). In multivariable random forest regression, BD-tau levels upon admission were most explained by stroke-related brain injury as assessed by final infarct volume, rather than baseline-related variables including age (Figure 2A). After admission, BD-tau showed a larger rise in patients with larger infarcts compared to those with smaller infarcts: the change of BD-tau in the first 24 hours after stroke onset was significantly associated with final infarct volume (β, 0.23 [95% CI, 0.19 to 0.28]; *P*<.0001) (Figure 2B). Similar patterns of the course of BD-tau over time were observed when restricting the analysis to patients with known symptom onset (N=362 [72%]; *P*<.0001) (eFigure 4 in the Supplement) and when stratifying patients with ischemic stroke for admission ASPECTS (eFigure 5 in the Supplement). The hourly rate of change of BD-tau between admission (day 1) and day 2 (ΔBD-tau) was higher in patients with lower collateral supply upon admission (*R*^2^=0.10, *P*<.0001) (Figure 2C), when adjusting for ischemic core volume (adjusted *R*^2^=0.10, *P*<.0001) and when using the more granular regional leptomeningeal collateral score (*R*^2^=0.12, *P*<.0001). ΔBD-tau was significantly correlated with infarct progression (*ρ*=0.58, *P*<.0001) (eFigure 6 in the Supplement) and almost exclusively explained by final infarct volume (eFigure 7 in the Supplement). For comparison, BD-tau showed a higher relative increase within the first 24 hours (mean, 160% [standard deviation [SD], 426%]) compared with NfL (mean, 60% [SD, 124%]; *P*<.0001) (eFigure 8 in the Supplement). At day 2 (median time from onset to sample collection: 22.7 hours [IQR, 19.5 to 25.3]), BD-tau levels were highly predictive of final infarct volume (*ρ*=0.59, *P*<.0001) (Figure 2D), also when restricting the analysis to patients with MRI-based infarct volumetry (N=288 [57%]; *ρ*=0.47, *P*<.0001) (eFigure 9 in the Supplement).

**Figure 2.**
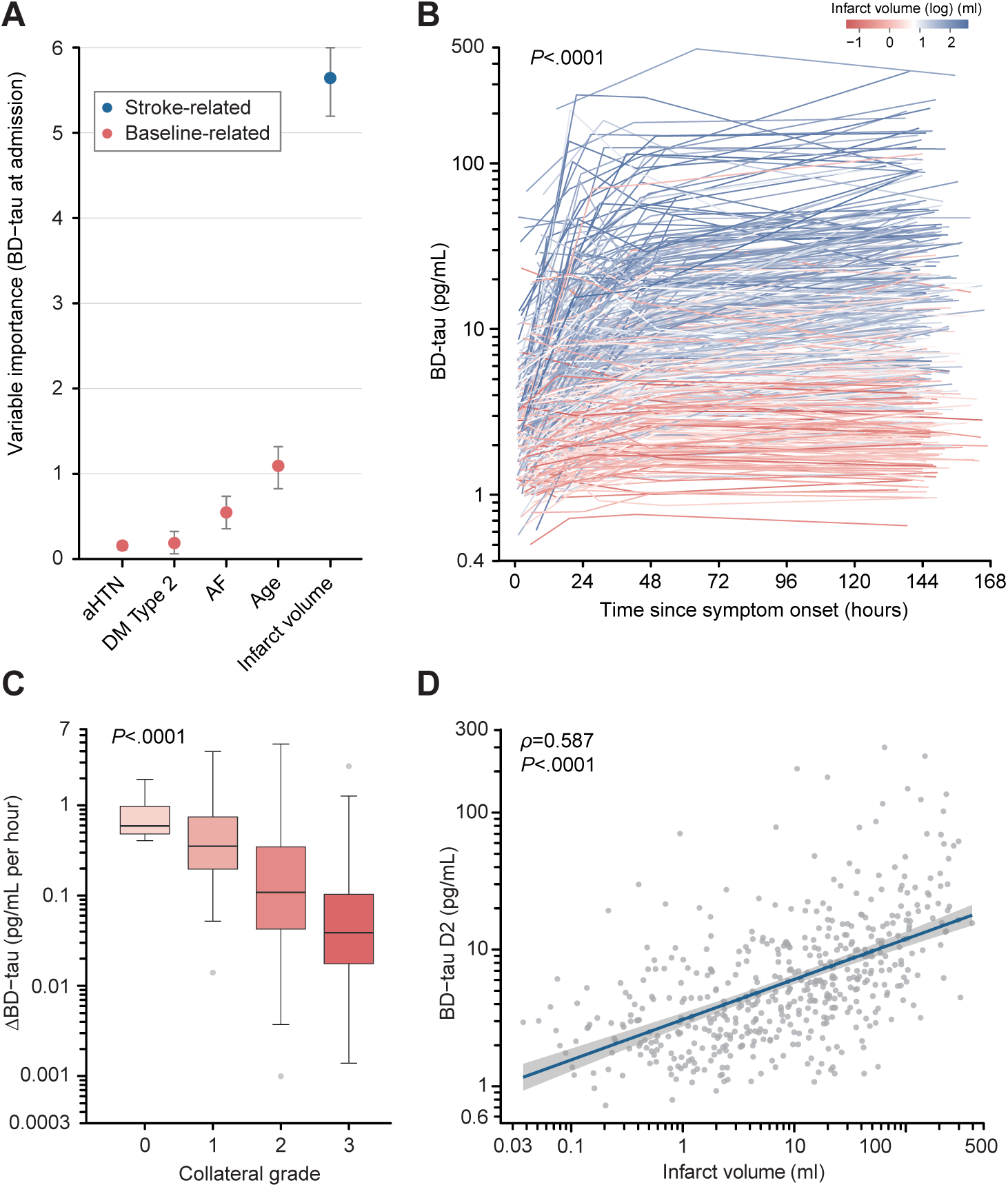
Relation of BD-tau with imaging-based metrics of brain injury. (**A**) Variable importance to predict BD-tau levels upon admission in a multivariable random forest regression model. Shown are the median and 5% and 95% quantiles of importance values. (**B**) Plasma levels of BD-tau are plotted over time in hours from stroke onset. Line color indicates final infarct volume. The *P* value was calculated for the effect of final infarct volume on the temporal course of BD-tau levels using a linear mixed model. (**C**) ΔBD-tau in relation to the degree of collateral supply. The collateral grade was assessed on CT angiography upon admission with a higher number indicating better collateral supply.^27^ The boxes represent (from bottom to top) the first quartile, the median and the third quartile. The upper whiskers indicate the third quartile plus 1.5 times the interquartile range while the lower whiskers indicate the first quartile minus 1.5 times the interquartile range. The *P* value was calculated using one-way analysis of variance. (**D**) BD-tau levels at day 2 in correlation with final infarct volume determined by delayed neuroimaging. The blue line indicates the linear fit and the grey area the 95% confidence interval. The *P* value was calculated using Spearman’s rank correlation. BD-tau, brain-derived tau. ΔBD-tau, change of BD-tau between admission and day 2 adjusted for the time passed in hours; CT, computerized tomography; aHTN, arterial hypertension; DM, diabetes mellitus; AF, atrial fibrillation.

### Relation of BD-tau to secondary events and recanalization

Following our findings on the relation of BD-tau with the extent of brain injury within the first 24 hours after stroke onset, we performed post-hoc analyses on the relation of longitudinal BD-tau changes with secondary events within the first seven days including intracerebral hemorrhage, early recurrent ischemic stroke, and hemorrhagic transformation: in analysis adjusted for final infarct volume, patients with either secondary intracerebral hemorrhage (Figure 3A), early recurrent ischemic stroke (Figure 3B), hemorrhagic transformation (Figure 3C), or specifically parenchymal hematoma (eFigure 10 in the Supplement) showed similar BD-tau levels upon admission but developed higher BD-tau levels over the next days compared with patients without any secondary event.

**Figure 3.**
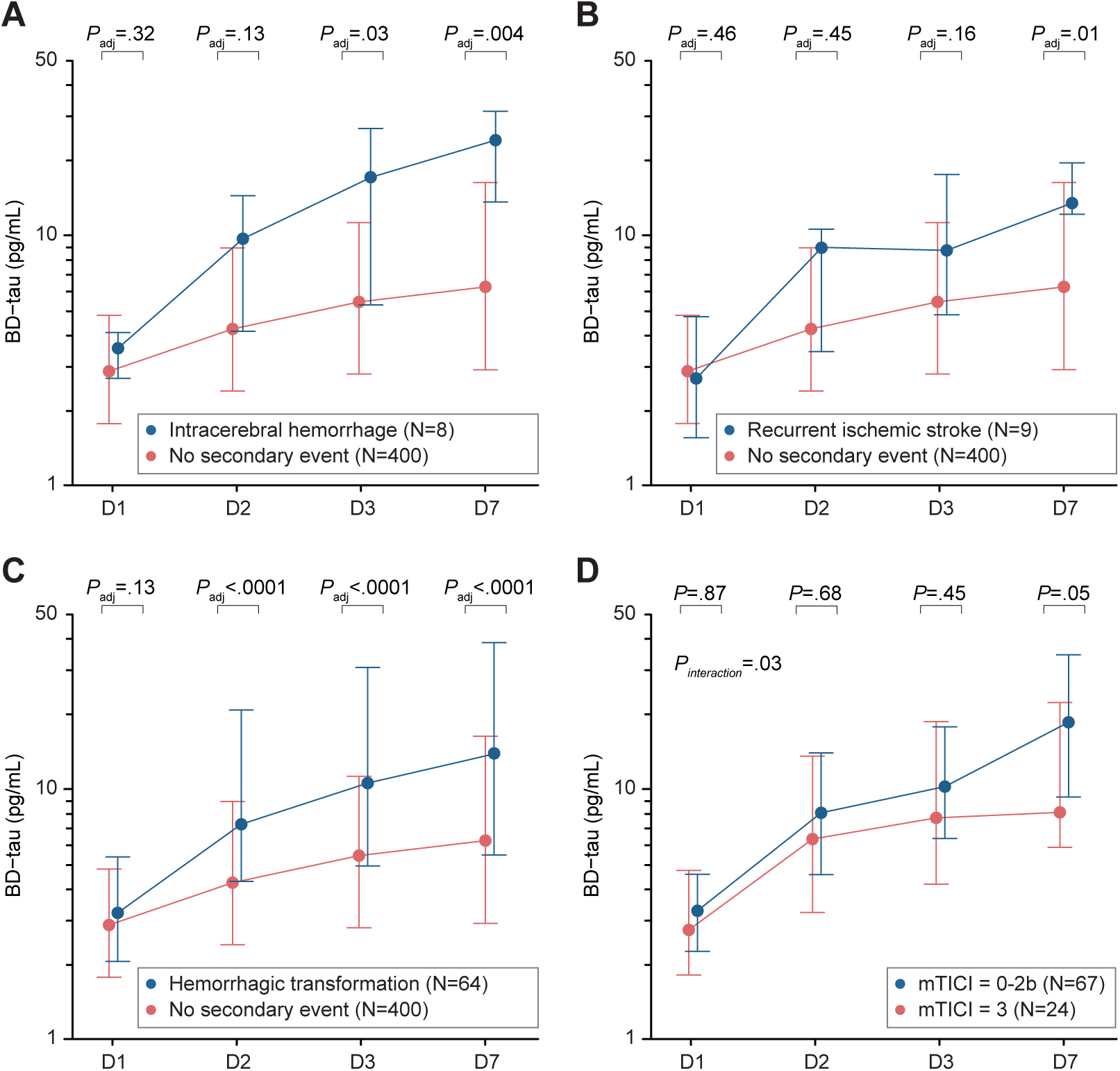
Relation of BD-tau with secondary events and recanalization. BD-tau levels upon admission (D1), at day 2, day 3, and day 7 in patients without any secondary event within the first seven days compared with (**A**) patients with secondary intracerebral hemorrhage, (**B**) patients with recurrent ischemic stroke, and (**C**) patients with hemorrhagic transformation of the index infarct. (**D**) BD-tau levels upon admission (D1), at day 2, day 3, and day 7 in patients with large-vessel occlusion stroke of the anterior circulation that underwent endovascular treatment with complete recanalization (mTICI=3) compared with patients with incomplete recanalization (mTICI=0-2b). Analysis restricted to patients without any secondary event during the next seven days. Groups were matched for BD-tau levels upon admission and occluded vessel site (eMethods in the Supplement). The dots represent the median values, the whiskers the first and third quartile. *P* values were calculated using an ANOVA, adjusted for infarct volume. BD-tau, brain-derived tau; IS, ischemic stroke; D1, day 1; mTICI, modified Thrombolysis In Cerebral Infarction.

In post-hoc analyses on the relation of BD-tau changes with the final grade of recanalization after endovascular treatment, patients with large-vessel occlusion stroke of the anterior circulation and incomplete recanalization (mTICI=0-2b) developed higher BD-tau levels over time compared with patients with complete recanalization (mTICI 3; *P_interaction_* =.03) (Figure 3D). In analysis restricted to patients with successful recanalization, BD-tau levels from admission to day 7 were significantly higher in patients with 90-day functional dependence compared with patients with functional independence (eFigure 11 in the Supplement).

### Relation of BD-tau to clinical severity and prediction of functional outcome after stroke

BD-tau upon admission was significantly associated with the NIHSS score upon admission (*ρ*=0.34, *P*<.0001) (Figure 4A) as was BD-tau at day 2 with the 24-hour NIHSS score (*ρ*=0.54, *P*<.0001) (eFigure 12 in the Supplement). ΔBD-tau was higher in patients with early neurological deterioration (*P*=.007) (Figure 4B). Higher BD-tau levels upon admission were associated with higher mRS scores (i.e. worse functional outcome) at 90 days (OR, 11.53; 95% CI, 6.17 to 21.55; *P*<.0001) (Figure 4C), also when adjusting for age, sex, hypertension, and premorbid mRS (adjusted OR [aOR], 5.54; 95% CI, 2.86 to 10.72; *P*<.0001). Similar results were found for the relation of BD-tau at day 2, day 3, and day 7 with mRS scores at 90 days (all *P*<.0001, eFigure 13 in the Supplement) and for the relation of BD-tau with mRS scores at day 7 (all *P*<.0001, eFigure 14 in the Supplement) as well as with the rates of functional independence at day 7 and day 90 (all *P*<.0001, eFigures 15 and 16 in the Supplement). In random forest regression, BD-tau at day 2 showed higher value in predicting 90-day functional outcome compared with infarct volume (Figure 4D), also when restricting the analysis to patients with MRI-based infarct volumetry (eFigure 17 in the Supplement).

**Figure 4.**
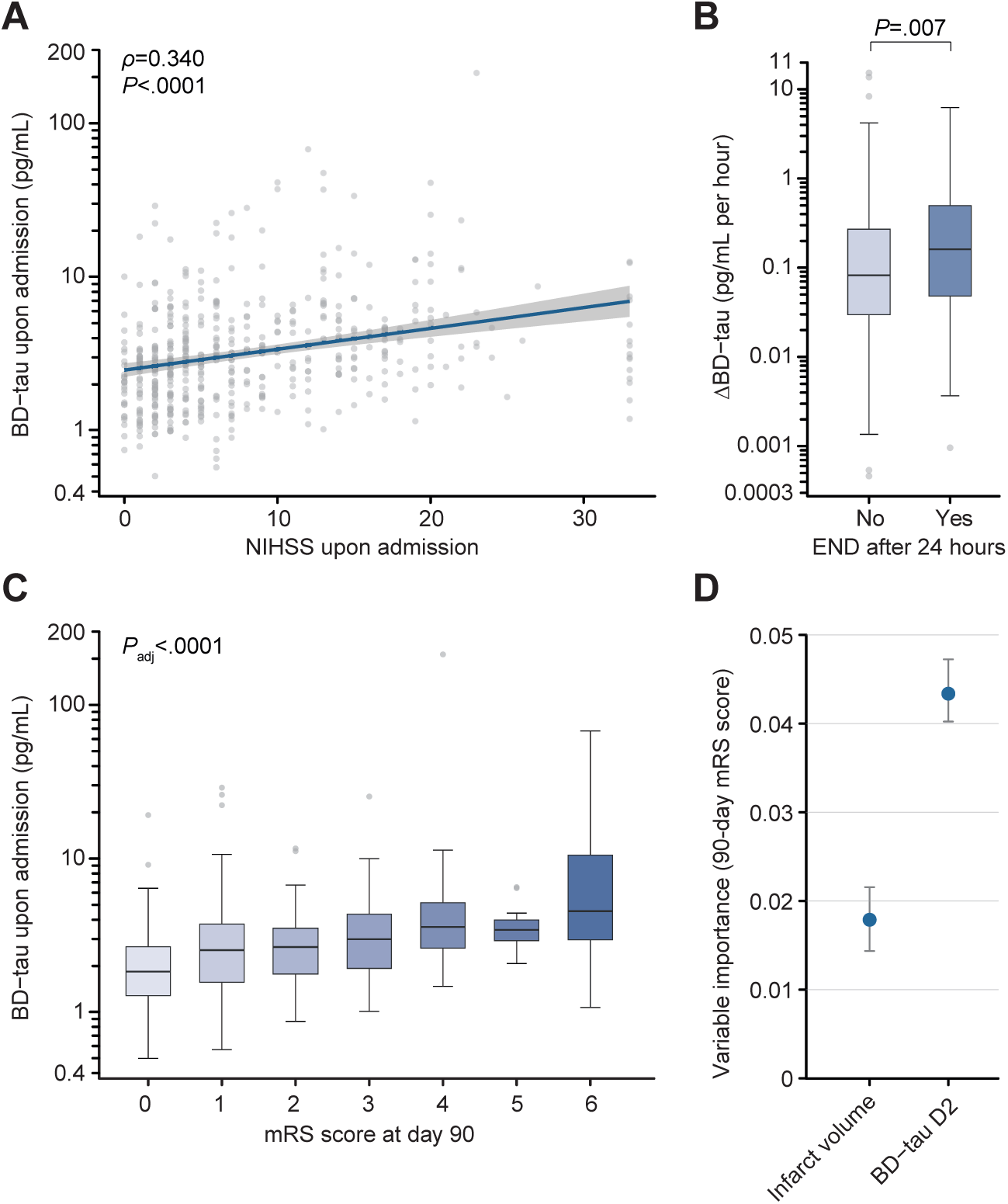
Relation of BD-tau with clinical severity and functional outcome. (**A**) BD-tau levels upon admission in relation to the NIHSS score upon admission. The blue line indicates the linear fit and the grey area the 95% confidence interval. The *P* value was calculated using Spearman’s rank correlation. (**B**) ΔBD-tau in patients with early neurological deterioration (END) compared with ΔBD-tau in patients without END. The boxes represent (from bottom to top) the first quartile, the median and the third quartile. The upper whiskers indicate the third quartile plus 1.5 times the interquartile range while the lower whiskers indicate the first quartile minus 1.5 times the interquartile range. The *P* value was calculated using an ANOVA. (**C**) BD-tau levels upon admission in relation to functional outcome at day 90 as assessed by the mRS score. The boxes represent (from bottom to top) the first quartile, the median and the third quartile. The upper whiskers indicate the third quartile plus 1.5 times the interquartile range while the lower whiskers indicate the first quartile minus 1.5 times the interquartile range. The *P* value was calculated using a multivariable logistic regression model adjusting for age, sex, hypertension, and the premorbid mRS score. (**D**) Variable importance to predict functional outcome at day 90 in a multivariable random forest regression model. Shown are the median and 5% and 95% quantiles of importance values. NIHSS, National Institutes of Health Stroke Scale; mRS, modified Rankin Scale; END, early neurological deterioration; ΔBD-tau, change of BD-tau between admission and day 2 adjusted for the time passed in hours; BD-tau, brain-derived tau.

## DISCUSSION

In a prospective cohort study of more than 500 IS patients, plasma BD-tau levels demonstrated suitability to monitor ongoing brain injury in acute IS: (i) BD-tau upon admission was significantly correlated with ASPECTS and predictive of final infarct volume, (ii) BD-tau increased within the first 24 hours in correlation with infarct progression, (iii) BD-tau at 24 hours was highly predictive of final infarct volume and was superior to infarct volume in predicting 90-day functional outcome, (iv) patients with secondary events demonstrated higher BD-tau levels compared with those without these events: with hemorrhagic transformation as early as day 2, intracerebral hemorrhage by day 3, and early recurrent stroke by day 7, and (v) patients with incomplete recanalization showed higher BD-tau levels by day 7 compared with patients with complete recanalization.

Our study identifies BD-tau as the potentially first blood-based biomarker that rapidly reflects ongoing brain injury in acute ischemic stroke. BD-tau levels and their dynamics over time might be particularly useful to (i) support clinical decision-making regarding endovascular treatment, e.g. to quantify lost tissue after inter-hospital transfer or to stratify patients presenting with low NIHSS scores according to BD-tau dynamics, (ii) identify ongoing brain injury in patients presenting beyond 24 hours^31^ who might still benefit from recanalization strategies,^6^ (iii) select patients with successful recanalization but ongoing brain injury for trials evaluating cytoprotection or targeting clinically ineffective reperfusion,^11,12,32,33^ and (iv) to diagnose stroke in settings with limited neuroimaging resources. High-frequency monitoring of the development of BD-tau levels over time might further allow to map infarct growth in individual patients and improve our understanding of human stroke pathophysiology by providing insights into the actual timing and extent of neuronal death.^34,35^ The presented findings additionally support the notion that monitoring BD-tau levels after IS might be of value to detect secondary events including intracerebral hemorrhage and recurrent ischemic stroke: patients with post-stroke neurological deterioration and increasing BD-tau levels might be selected for neuroimaging rather than those with constant BD-tau levels. Similarly, tracking BD-tau over time might inform on ongoing brain injury and ischemic events in patients with subarachnoid hemorrhage, vasoconstriction syndromes, and cerebral vasculitis. Our findings on the superior value of 24-hour BD-tau levels to predict 90-day functional outcome compared with imaging-based infarct volume emphasize the potential of BD-tau to serve as a surrogate marker for the efficacy of treatments including of reperfusion strategies. Data from clinical trials are necessary to provide further evidence and insights into this. Conceptually, these findings could reflect that circulating brain-derived molecules detected by single-molecule assays might be able to capture more nuances of stroke brain injury, including selective neuronal loss,^36^ compared with neuroimaging. BD-tau might thus be more precise in detecting and monitoring the extent of brain injury compared with imaging-based infarct volumetry, which was recently found to explain only 12% of the treatment effect from endovascular treatment.^37^

Previously studied biomarkers for brain injury, such as NfL,^13^ GFAP,^15,16^ NSE,^14^ and S100B,^17^ either lacked specificity for the brain or failed to gauge the extent of brain injury sufficiently early. BD-tau addresses both requirements. Firstly, its assay was engineered to specifically capture tau emanating from the CNS^18^ in contrast to assays measuring total tau, which encompasses BD-tau and tau from peripheral tissues.^38,39^ Secondly, BD-tau elevated within hours after stroke, rising earlier than NfL, in our cohort. This observation aligns with prior data on the delayed response of NfL to brain injury^13^ and its earlier detection by BD-tau.^24^ It remains speculative to assume that this difference might be rooted in the dynamic interaction of tau with microtubules^20,21^ primarily in dendrites^40,41^ as opposed to NfL’s more static role as a core filament in axons.^42^ Notably, while axons start to fully degrade only 72 hours after injury,^23^ dendrites are among the first structural components to retract and degrade upon ischemia.^22^

Current diagnostic algorithms to assess brain injury in stroke primarily rely on neuroimaging techniques such as non-contrast imaging, CT perfusion, and MRI. While offering detailed insights into brain structures, their limitations include restricted availability in both resource-limited and well-equipped settings,^43,44^ unsuitability for certain patients, challenges associated with logistics including an elevated risk for complications in severely ill patients, high costs, and potential radiation exposure. Consequently, they may not be ideal for frequent monitoring of brain injury. Blood-based biomarkers, in contrast, are less invasive and have the potential for wider accessibility, particularly when assessed at the point-of-care. The adaptability of blood BD-tau levels for regular monitoring could make the assessment of stroke even more precise, complementing existing neuroimaging-based algorithms.

## LIMITATIONS

Our study has several limitations. The observational design has inherent biases including by confounding, by selection bias, and by time-related biases such as survivorship bias. Though our sample size was robust, the cohort was recruited at a single center and does not capture the full diversity of the global stroke patient population. As such, validation of our results in other cohorts, ideally in multi-center studies that span different countries and continents, is needed before they can be extrapolated. Our study was not designed to detect how informative actual BD-tau changes between two separate measurements 30 or 60 minutes apart are. Larger studies are needed to establish reference ranges, both of absolute values and dynamic changes, before BD-tau levels can be used to inform clinical management. Lastly, our study is centered on ischemic stroke and cannot inform on the value of BD-tau in hemorrhagic stroke.

## CONCLUSIONS

In 502 patients with acute IS, plasma BD-tau was associated with imaging-based metrics of brain injury upon admission, increased within the first 24 hours in correlation with infarct progression, and at 24 hours, was highly predictive of final infarct volume and superior to final infarct volume in predicting the 90-day modified Rankin Scale score. Further research is needed to validate the findings in diverse populations and to determine the potential role of plasma BD-tau in stroke care.

## Data Availability

The data produced in the present study are currently not available if not already presented in the manuscript.

## POTENTIAL CONFLICTS OF INTERESTS

KB has served as a consultant and at advisory boards for Acumen, ALZPath, BioArctic, Biogen, Eisai, Lilly, Moleac Pte. Ltd, Novartis, Ono Pharma, Prothena, Roche Diagnostics, and Siemens Healthineers; has served at data monitoring committees for Julius Clinical and Novartis; has given lectures, produced educational materials and participated in educational programs for AC Immune, Biogen, Celdara Medical, Eisai and Roche Diagnostics; and is a co-founder of Brain Biomarker Solutions in Gothenburg AB (BBS), which is a part of the GU Ventures Incubator Program, outside the work presented in this paper. TL protors and consults for Stryker, phenox, Acandis, and has in the past received suport for travel and service related fees from Medtronic, Pfizer, Cerus Endovascular, Sequent, and Microvention. NV, NLK, MD, and ST report a patent on the use of BD-tau. No other disclosures were reported.

## FUNDING

F.G.-O. was funded by the Anna Lisa and Brother Björnsson’s Foundation and Emil och Maria Palms Foundation. S. T. was supported by grants from the Corona foundation (S199/10081/2020), the Leducq foundation (21CVD04), and the Friedrich-Baur-Stiftung (52/23). TKK was supported by grants P30 AG066468, RF1 AG052525-01A1, R01 AG053952-05, R37 AG023651-17, RF1 AG025516-12A1, R01 AG073267-02, R01 AG075336-01, R01 AG072641-02, and P01 AG025204-16 from the National Institutes of Health; the Swedish Research Council (Vetenskåpradet; 2021-03244); the Alzheimer’s Association (AARF-21-850325); the Swedish Alzheimer Foundation (Alzheimerfonden); the Aina (Ann) Wallströms and Mary-Ann Sjöbloms stiftelsen; and the Emil och Wera Cornells stiftelsen. KB is supported by the Swedish Research Council (#2017-00915 and #2022-00732), Hjärnfonden, Sweden (#FO2017-0243 and #ALZ2022-0006), and the Swedish state under the agreement between the Swedish government and the County Councils, the ALF-agreement (#ALFGBG-715986 and #ALFGBG-965240).

## Notes

### Funding Statement

F.G.-O. was funded by the Anna Lisa and Brother Bjornssons Foundation and Emil och Maria Palms Foundation. S. T. was supported by grants from the Corona foundation (S199/10081/2020), the Leducq foundation (21CVD04), and the Friedrich-Baur-Stiftung (52/23). TKK was supported by grants P30 AG066468, RF1 AG052525-01A1, R01 AG053952-05, R37 AG023651-17, RF1 AG025516-12A1, R01 AG073267-02, R01 AG075336-01, R01 AG072641-02, and P01 AG025204-16 from the National Institutes of Health; the Swedish Research Council (Vetenskapradet; 2021-03244); the Alzheimers Association (AARF-21-850325); the Swedish Alzheimer Foundation (Alzheimerfonden); the Aina (Ann) Wallstroms and Mary-Ann Sjobloms stiftelsen; and the Emil och Wera Cornells stiftelsen. KB is supported by the Swedish Research Council (#2017-00915 and #2022-00732), Hjarnfonden, Sweden (#FO2017-0243 and #ALZ2022-0006), and the Swedish state under the agreement between the Swedish government and the County Councils, the ALF-agreement (#ALFGBG-715986 and #ALFGBG-965240).

### Author Declarations

Ethic committee of LMU Munich gave ethical approval for this work.

