## Supplemental methods and data for "Brain-derived Tau for Monitoring Brain Injury in Acute Ischemic Stroke"

### 1 eMethods

**Participant screening and procedures intended to ensure a representative sample of eligible patients**

Patients were identified by two independent and partially overlapping mechanisms: First, during working hours, research personnel continuously screened the hospital's clinical database for patients admitted with acute stroke-like symptoms to the emergency department. Second, hospital staff informed research personnel both during working hours and non-working hours of eligible patients admitted with acute stroke-like symptoms to the emergency department. Thus, recruitment occurred continuously, 24 hours a day, seven days a week. Further, screening procedures covered both patients directly admitted to LMU University Hospital and patients referred from a primary stroke center. To prevent a bias towards mildly affected patients, a deferred consent process was used for those initially unable to provide consent and when a legally authorized representative was not available (see section on deferred consent).

**Inclusion and exclusion criteria**

The final diagnosis of ischemic stroke was based on the presence of a diffusion weighted imaging (DWI)-positive lesion on MRI or a new lesion on a delayed CT scan.[1] Patients without a DWI-positive lesion on MRI or a new lesion on a delayed CT scan were classified as having stroke mimics.[1] Patients were excluded if they had no follow-up CT or MRI imaging, underwent major surgery or experienced myocardial infarction, ischemic stroke, transient ischemic attacks, traumatic brain injury, cerebral venous sinus thrombosis, any intracranial hemorrhage, thrombosis, or pulmonary embolism within the four weeks prior to admission, suffered from an in-house stroke, were comorbid for chronic inflammatory bowel disease, or had undergone percutaneous endoscopic gastrostomy. Patients could only be enrolled once in the study. Among initially eligible patients, we selected those who had blood samples collected upon admission (day 1), the next morning (day 2), day 3, and day 7 (or discharge if earlier).

**Collection and management of data including of functional outcome at follow-up**

During patients' hospitalization, research personnel reviewed patients' hospital charts and ensured the collection of blood samples. For unwitnessed onset stroke, time of onset was considered to be the midpoint between last seen well and time of recognition.[2] Clinical severity was assessed upon admission and at 24 hours using the National Institutes of Health Stroke Scale (NIHSS) score. Early neurological deterioration (END) was defined as any worsening in the NIHSS score from the assessment upon admission to the assessment at 24 hours. The diagnosis of an early recurrent ischemic stroke was based on i) sudden onset of a new focal neurological deficit within the first seven days after the index stroke that could not be explained by other non-ischemic causes such as seizure, metabolic derangement, or other systemic conditions, and ii) the presence of a new DWI-positive lesion on MRI or a new ischemic lesion on a delayed CT scan that was distinct from the index stroke lesion and that was consistent with the new clinical symptoms. Other adverse events that were registered were intracranial hemorrhages (N=20), cardiac arrest (N=2), and craniectomy (N=6). Functional outcome of patients with ischemic stroke was evaluated at day 7 (or discharge if earlier) and at day 90 using the modified Rankin Scale score ranging from 0 (no symptoms) to 6 (death). At day 90, the mRS was assessed either by face-to-face or telephone interview. mRS scores were assessed by trained neurologists. All clinical information of recruited patients was assessed independently by two members of the study team and finally reviewed by the investigator.

**Recruitment of healthy controls**

Healthy controls (HC) were recruited through a single outpatient clinic at LMU University Hospital. These were mostly spouses or companions of patients of the outpatient clinic. Compared to the recruited cohort of patients with ischemic stroke (IS), this cohort (N=247) comprised more women (HC: 156 women [63%] vs. IS: 208 women [41%], $P<0.0001$) and was younger (HC: median age [IQR]: 68 years [55-75] vs IS: 76 [66-83], $P<0.0001$). Selecting those healthy controls with an age of 66 years or higher (N=102), largely eliminated those differences (sex: HC: 44% vs. IS: 41%, P=0.66; age: HC: 75 years (71-78) vs. IS: 76 [66-83], P=0.84).

**Deferred consent**
A deferred consent process was used for those patients unable to provide consent (e.g. patients with aphasia) and when a legally authorized representative was not available, allowing collection of plasma samples while awaiting consent.

**Blood sampling and processing**
Blood samples were collected upon hospital admission in the emergency department (day 1), at the next morning (day 2), at day 3, and at day 7. Additional samples were collected at day 90 during follow-up for a subset of patients with ischemic stroke. Blood samples at day 2, 3, and 7 were collected in the morning in parallel to routine blood tests if possible. Whole blood was drawn into EDTA-plasma containers (Sarstedt) by venipuncture or from existing arterial or venous lines. After 30 to 45 minutes at room temperature, separation of plasma was achieved by differential centrifugation at 2000g for 10 minutes at 15 °C. Samples were aliquoted in screw cap vials and kept at -80 °C.

**BD-tau and NfL measurements**
Plasma BD-tau was quantified on the Simoa HD-X platform (Quanterix) using a previously described protocol.[3] In short, we used the sheep monoclonal antibody TauJ.5H3 (Bioventix Plc, Surrey, UK) for capture and an N-terminal-tau mouse monoclonal antibody for detection. Recombinant full-length tau-441 (TO8-50FN, SignalChem) was used as calibrator. Assay development and validation procedures, including dilution linearity and spike recovery, have recently been described.[3] Plasma Neurofilament Light Chain (NfL) was measured on the Simoa HD-X platform using a Quanterix assay (2-Plex B Lot #502247). Dilution factor for both markers was one in four. Six samples were diluted one in ten to fall within the range of the standard curve. Quality controls were run at the beginning and end of each plate. Experimenters were blinded for patient characteristics including group allocation and imaging-based metrics of brain injury.

**Neuroimaging studies and assessment of brain injury**
Multimodal CT was obtained as part of clinical routine upon hospital admission using a standard protocol that included noncontrast CT, CT angiography, and CT perfusion. CT examinations were performed on SOMATOM Definition Force, AS + and Flash scanners (Siemens Healthineers, Forchheim, Germany). CT perfusion data were processed using syngo Neuro Perfusion CT (Siemens Healthineers, Forchheim, Germany) including threshold-based calculation of the ischemic core (cerebral blood volume <1.2 mL/100 mL).[4] The Alberta Stroke Program Early CT Score (ASPECTS) on non-contrast CT[5] and collateral supply on CT angiography[6,7] were assessed by experienced radiologists. Data on ASPECTS were available for 459 patients (91.4%), data on collateral supply were available for 347 patients (69.1%), and data on ischemic core volume were available for 220 patients (43.8%). Final infarct volume was quantified on images from delayed diagnostic scans (at least 48 hours after stroke onset, mean time from onset to imaging: 4 days), either CT or MRI (diffusion-weighted, T2 or fluid-attenuated inversion recovery). The modality and image with the largest infarct size was used for volumetry. Trained raters segmented infarcts manually slice-by-slice. Infarct progression was calculated by subtracting ischemic core volume quantified on admission CT perfusion scans from final infarct volume quantified on delayed neuroimaging. Successful recanalization was defined as a final modified Thrombolysis in Cerebral Infarction (mTICI) score of 2b or 3. The occurrence of hemorrhagic transformation was evaluated based on the morphological ECASS criteria[8] by experienced radiologists. All raters were blinded for BD-tau levels.

**Further details of statistical analyses**
Differences in patient characteristics were assessed using the Kruskal-Wallis rank sum test, Pearson's Chi-squared test and the Fisher's exact test. To test for normality, we used the Shapiro-Wilk test. If significant, log transformation (base e) was used. Linear mixed models were used to test the temporal course of BD-tau: first to test differences between the different time points (fixed effect: days, random effect: patient ID) and second to test the interaction between hours since symptom onset and infarct volume (fixed effects) with patient ID as random effect. While plots displaying ΔBD-tau values are restricted to positive values on the log-scaled axis, statistical analyses include all available values. Matching was performed using the MatchIt package[9] using the full optimal matching method with a probit link, which assigns every treated and control unit in the sample to a subclass. Balance was improved after matching. To assess conditional variable importance in multivariable models we used random forest regression with standard parameters, grew 1,501 trees per run, and performed 100 runs to calculate 95% confidence intervals ('party' package in R).[10] For all analyses, complete case

analysis was performed without imputations. All analyses were performed in ‘R’, version 4.2.2.

**Serial high-frequency sampling to assess real-time evolution of BD-tau after stroke onset**
To capture the real-time evolution of BD-tau levels within the first 36 hours after onset, we sampled two patients with ischemic stroke with high frequency from admission (three and five hours after onset) until at least 36 hours after onset. Legally authorized representatives gave informed consent in accord with ethical approval (Ref. No. 121-09). Plasma samples were collected hourly, except between 3AM and 5AM in the morning and during diagnostic procedures. Plasma samples were processed as described in the eMethods section ‘Blood sampling and processing’.

### 2 eFigures

#### eFigure 1. Concentrations of plasma BD-tau in internal quality controls

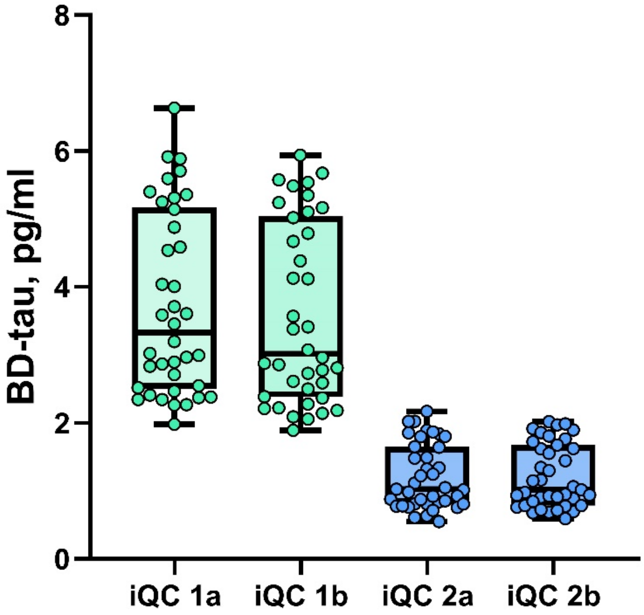

Variation intra and inter-runs was assessed using two internal quality controls (iQC) (1 and 2) at the beginning (a) and the end of each plate (b). Mean concentration for iQC 1 (n=76) was 3.7±1.32 pg/ml with a CV of 8.5% in the whole cohort. Mean concentration for iQC 2 (n=78) was 1.2±0.46 with a CV of 9.1% in the whole cohort. CV, coefficient of variation.

### eFigure 2. Study flowchart

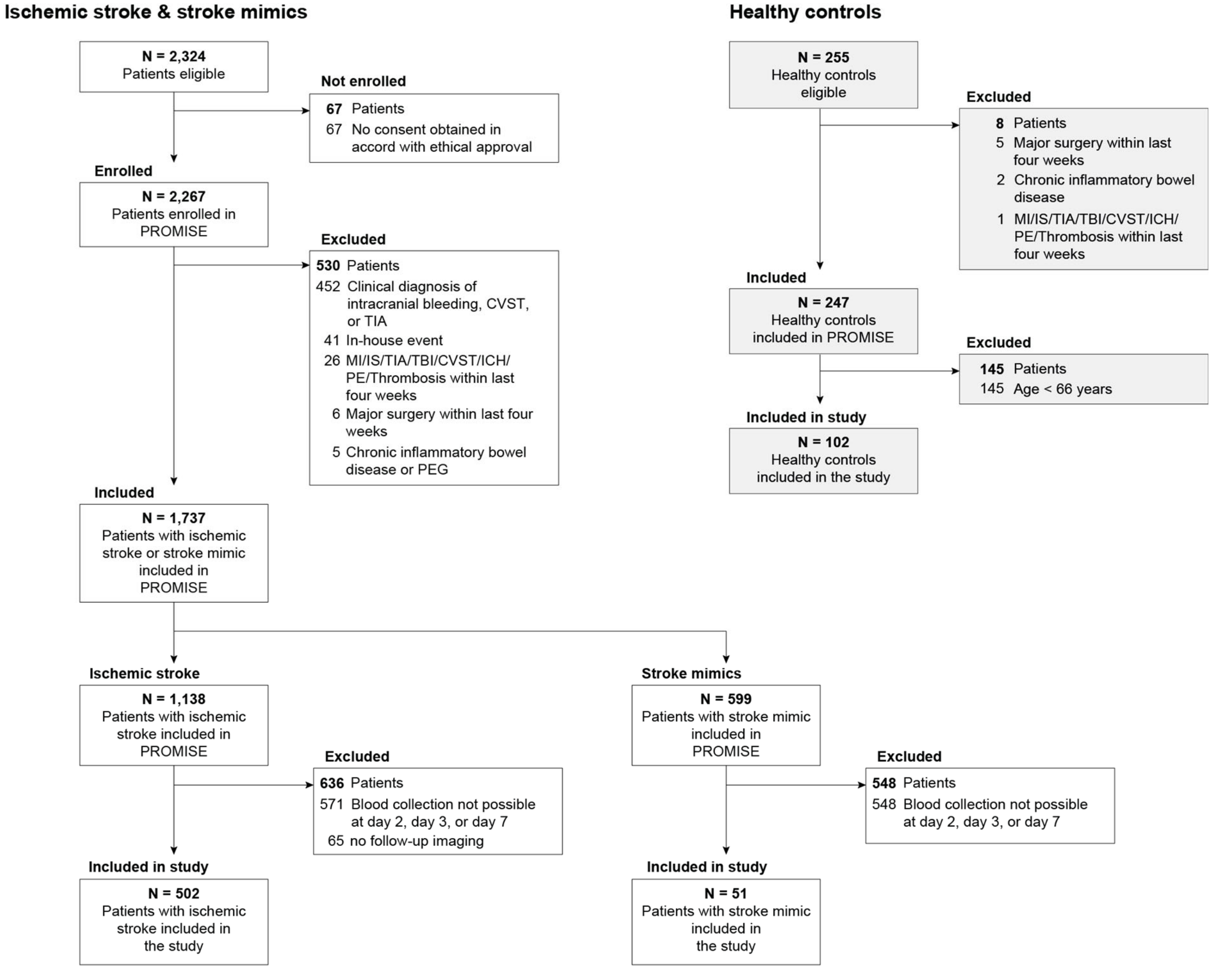

Flowchart of included patients in PROMISE stratified by diagnostic group. MI, myocardial infarction; IS, ischemic stroke; TIA, transient ischemic attack; CVST, cerebral venous and sinus thrombosis; TBI, traumatic brain injury; ICH, intracranial hemorrhage; PE, pulmonary embolism; PEG, percutaneous endoscopic gastrostomy.

### eFigure 3. Correlation of BD-tau upon admission with final infarct volume.

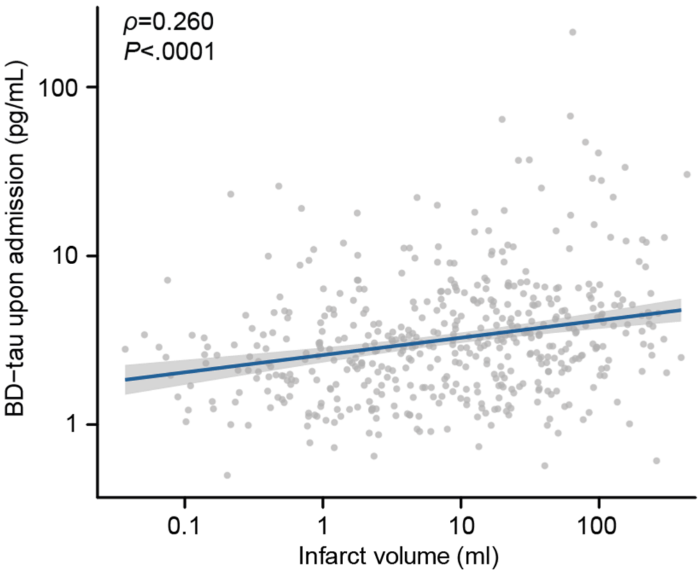

BD-tau levels upon admission in relation to final infarct volume determined by delayed neuroimaging. The P value was calculated using Spearman's rank correlation. The blue line indicates the linear fit. The grey area indicates the 95% confidence interval. BD-tau, brain-derived tau.

**eFigure 4. Development of BD-tau levels over time in stroke patients with known symptom onset.**

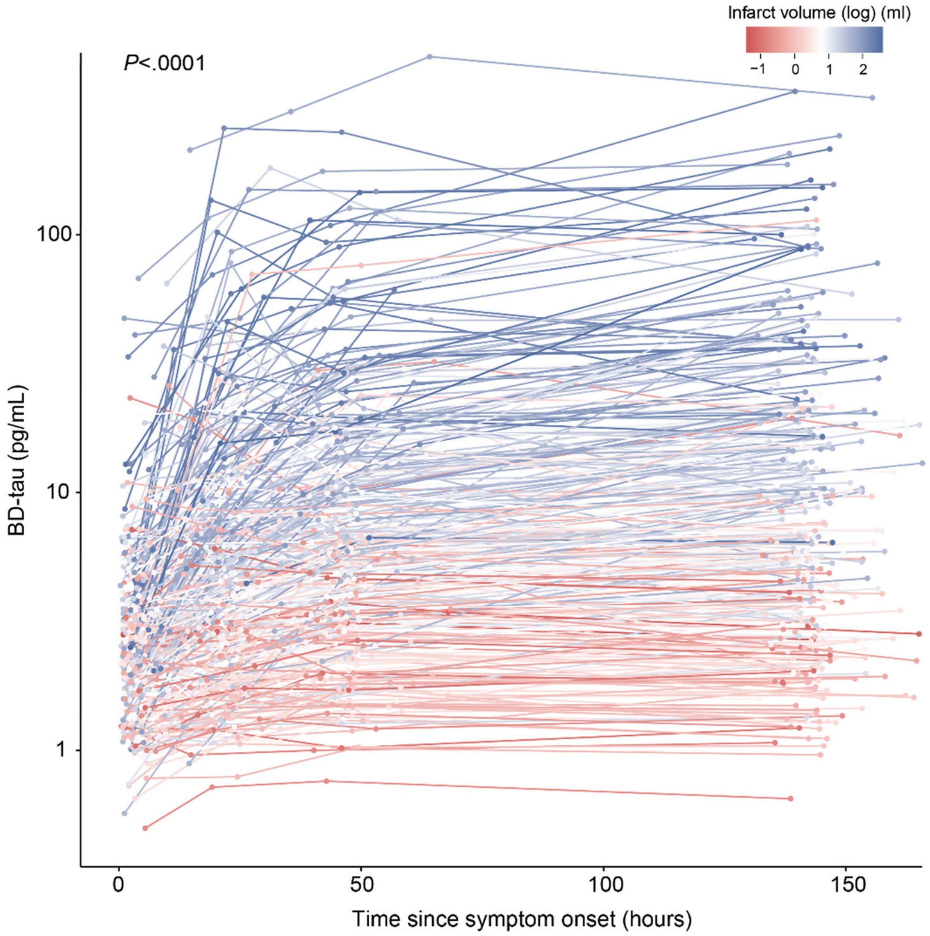

Shown is the development of Brain Derived-tau (BD-tau) over time in hours since onset in those patients with ischemic stroke where the time of onset was known. Each line represents an individual patient with ischemic stroke. Line color indicates final infarct volume. The P-value was calculated for the effect of infarct volume on the temporal course of BD-tau levels using a linear mixed model.

### eFigure 5. Development of BD-tau levels over time after stroke stratified by ASPECTS

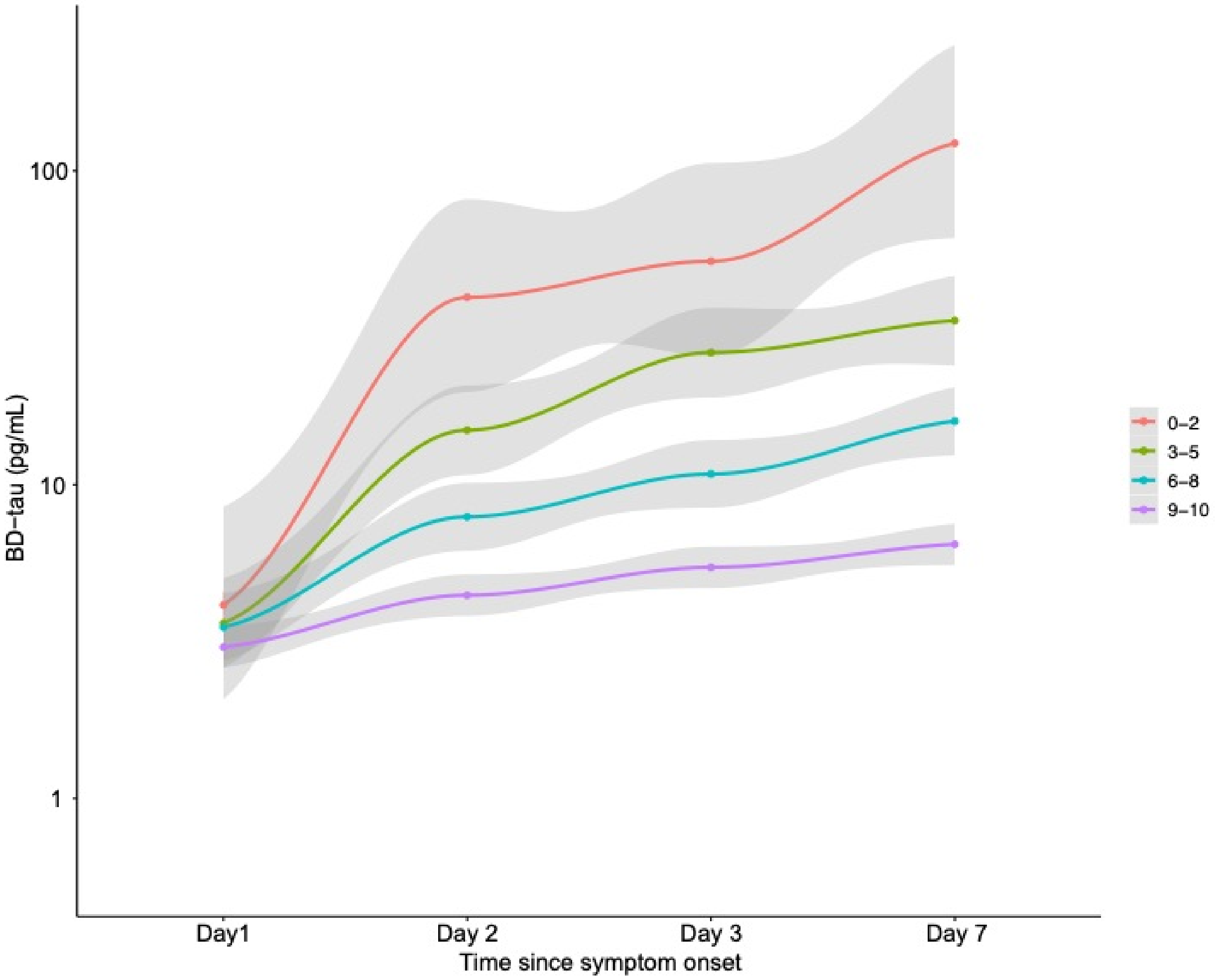

Shown is the development of Brain Derived-tau (BD-tau) over days since symptom onset stratified by ASPECTS upon admission. A higher ASPECTS represents less severe stroke severity on admission CT. Grey areas indicate 95% confidence intervals. BD-tau, brain-derived tau; ASPECTS, Alberta Stroke Program Early CT Score; CT, computerized tomography.

### eFigure 6. Correlation of ΔBD-tau with infarct progression

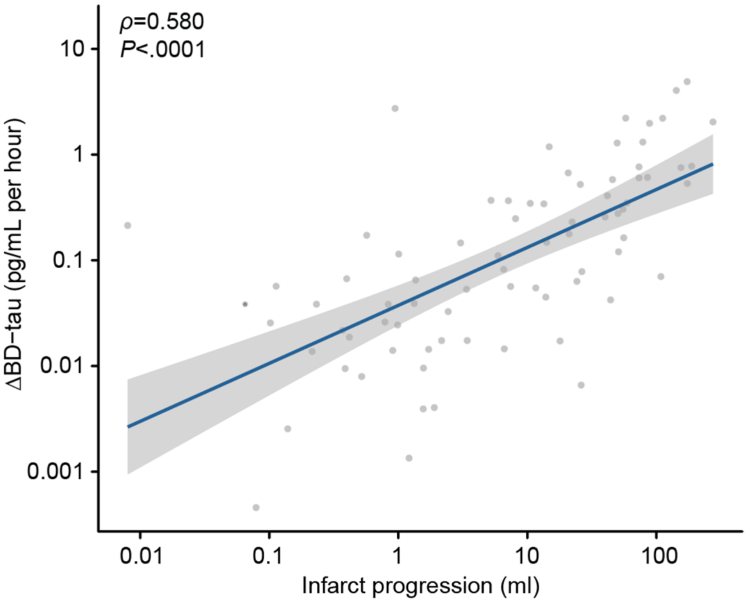

ΔBD-tau in relation to infarct progression. The P value was calculated using Spearman's rank correlation. The blue line indicates the linear fit. The grey area indicates the 95% confidence interval. Infarct progression was calculated by subtracting ischemic core volume quantified on admission CT perfusion from infarct volume quantified on delayed neuroimaging. BD-tau, brain-derived tau; CT, computed tomography.

### eFigure 7. Variable importance for ΔBD-tau.

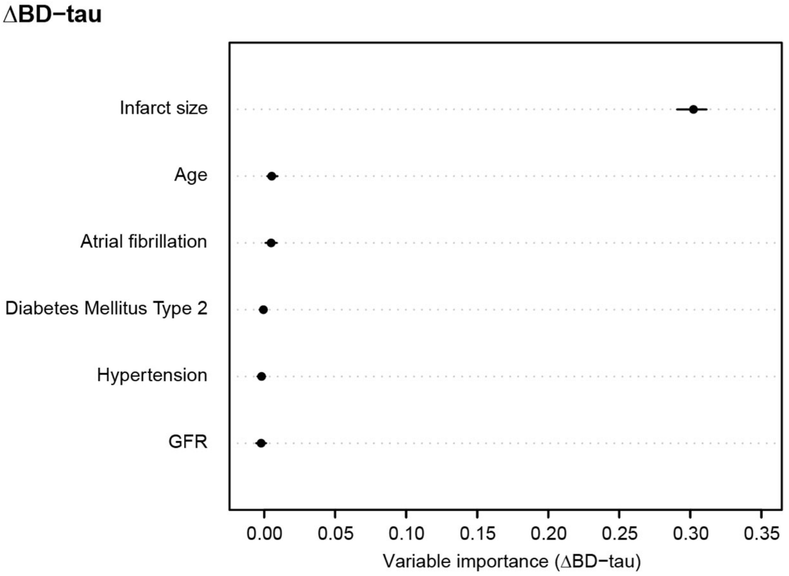

Shown are the median (dot) and 5% and 95% quantiles (whiskers) of importance values to predict ΔBD-tau in a multivariable random forest regression model. BD-tau, brain-derived tau; GFR, glomerular filtration rate.

### eFigure 8. Development of NfL levels over time after stroke onset

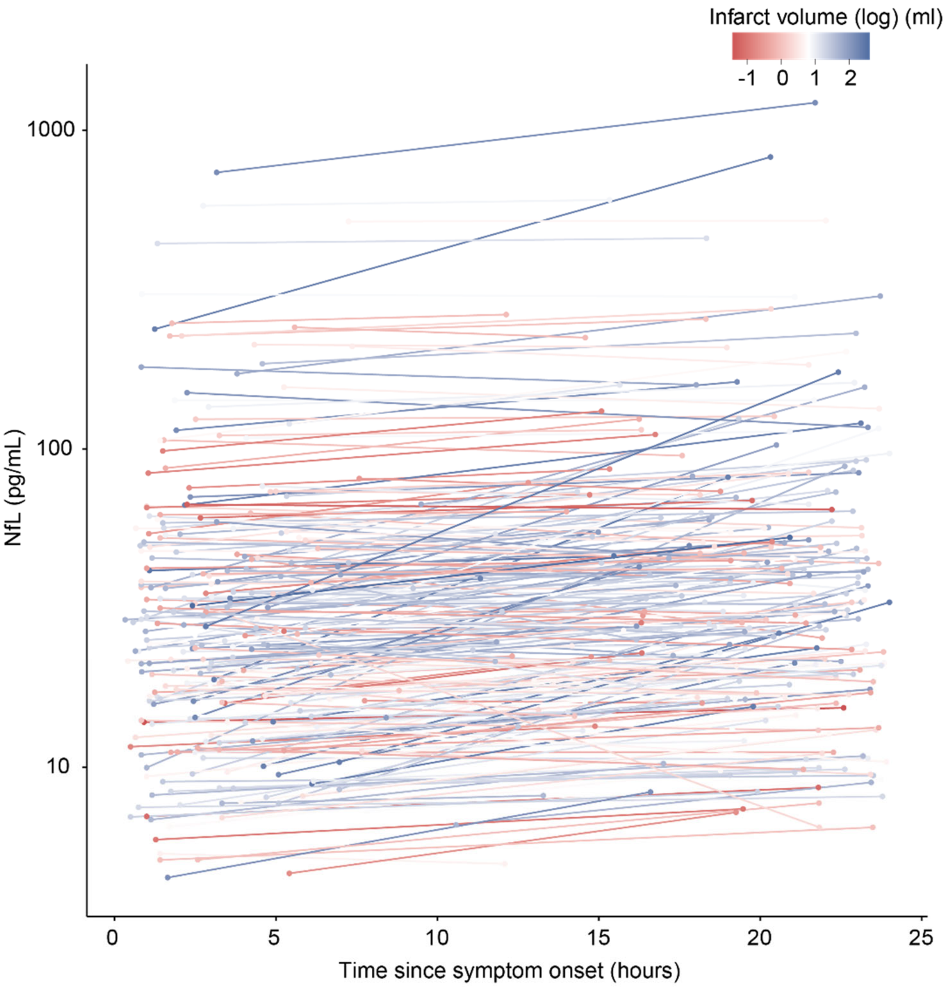

Shown is the development of Neurofilament Light chain (NfL) over time within the first 24 hours after stroke onset. Each line represents an individual patient with ischemic stroke. Line color indicates infarct size. NfL, Neurofilament Light chain.

**eFigure 9. Correlation of BD-tau at day 2 with final infarct volume in analysis restricted to patients with MRI-based infarct volumetry.**

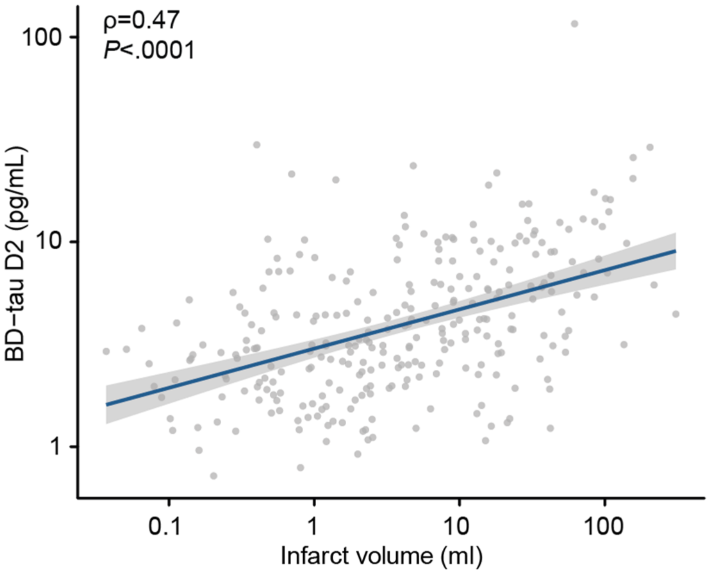

BD-tau levels at day 2 in relation to final infarct volume determined by delayed neuroimaging when restricting the analysis to patients with MRI-based infarct volumetry (N=288, 57%). The P value was calculated using Spearman's rank correlation. The blue line indicates the linear fit. The grey area indicates the 95% confidence interval. BD-tau, brain-derived tau; MRI, magnetic resonance imaging.

### eFigure 10. BD-tau in relation to parenchymal hematoma

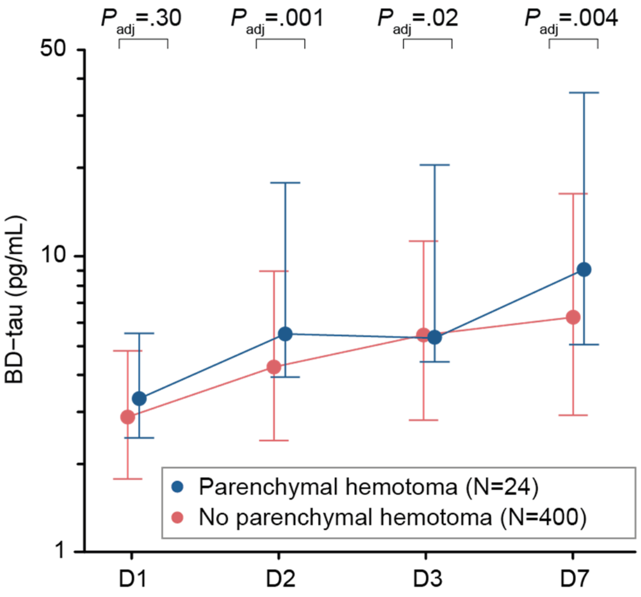

Shown are the median (dot) and interquartile range (whiskers) of BD-tau levels in patients with parenchymal hematoma (PH1 or 2 according to the ECASS classification[8]) compared to patients without parenchymal hematoma or any other secondary event. *P* values were calculated using an ANOVA adjusted for final infarct volume. BD-tau, brain-derived tau; PH, parenchymal hematoma; ANOVA, analysis of variance.

**eFigure 11. BD-tau levels in patients with successful recanalization after endovascular treatment stratified for the 90-day status of functional independence**

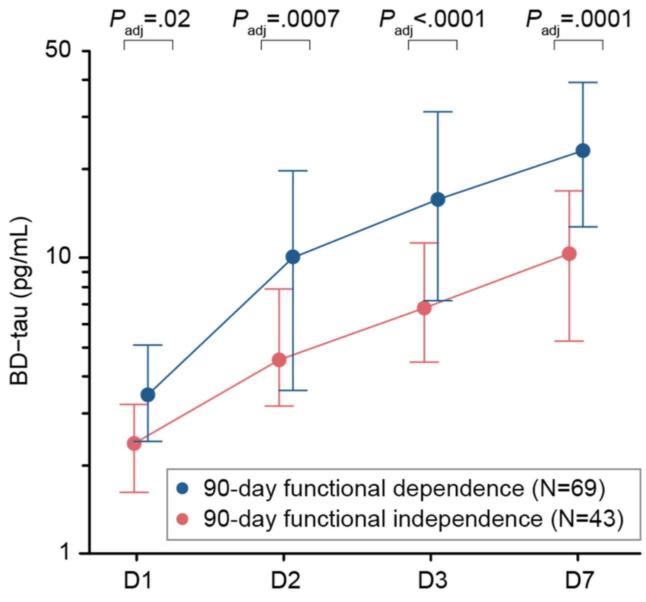

Shown are the median (dot) and interquartile range (whiskers) of BD-tau levels in patients with successful recanalization after endovascular treatment. Successful recanalization was defined as a final mTICI score of 2b or 3. *P* values were calculated using a multivariable logistic regression model adjusting for age, sex, hypertension, and the premorbid mRS score. BD-tau, brain-derived tau; mRS, modified Rankin Scale; MRI, magnetic resonance imaging; mTICI, modified Thrombolysis In Cerebral Infarction.

**eFigure 12. Correlation of BD-tau at day 2 with 24-hour NIHSS score.**

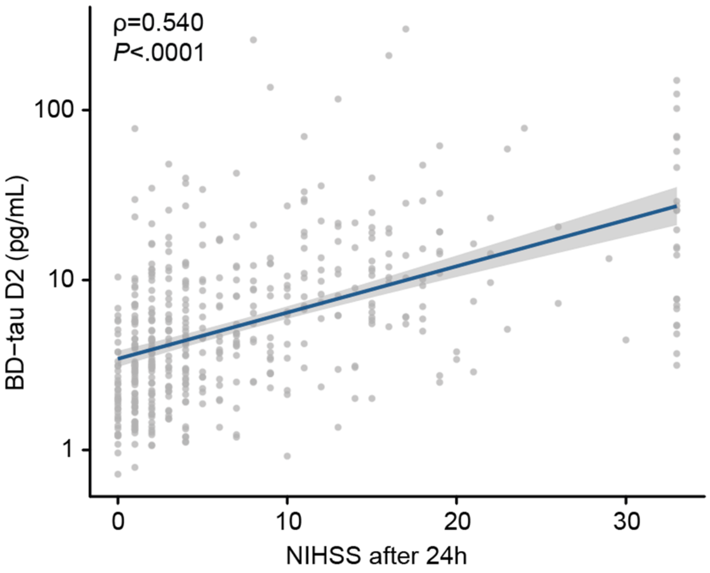

Shown is the association between BD-tau levels at day 2 and the 24-hour NIHSS score. The P value was calculated using spearman correlation. The linear fit is shown in blue. The 95% confidence interval is shown in grey. BD-tau, brain-derived tau; NIHSS, National Institutes of Health Stroke Scale.

**eFigure 13. Association of BD-tau upon admission, at day 2, day 3, and day 7 with mRS scores at 90 days.**

| BD-tau sampling | aOR (95% CI) |
|---|---|
| Upon admission | 5.54 (2.86-10.72) |
| Day 2 | 7.28 (4.36-12.14) |
| Day 3 | 7.76 (4.78-12.59) |
| Day 7 | 6.87 (4.51-10.48) |

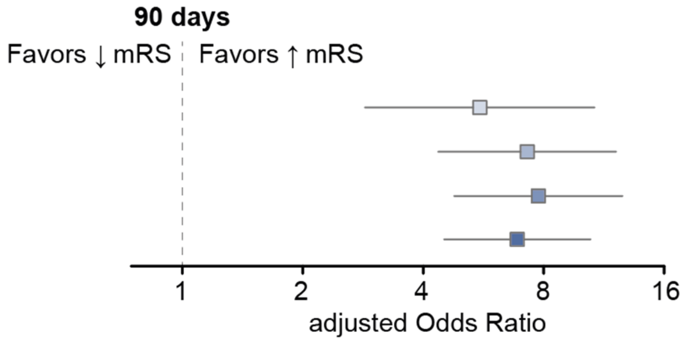

Shown is a forest plot of the associations of BD-tau levels upon admission, at day 2, day 3, and day 7 with mRS scores at 90 days. The odds ratios (95% confidence interval) were calculated using a multivariable logistic regression model adjusting for age, sex, hypertension, and the premorbid mRS score. mRS, modified Rankin Scale; aOR, adjusted Odds ratio; CI, Confidence Interval; BD-tau, brain-derived tau.

**eFigure 14. Association of BD-tau over time after stroke with mRS scores at day 7.**

| BD-tau sampling | aOR (95% CI) |
|---|---|
| Upon admission | 4.75 (2.76-8.16) |
| Day 2 | 8.79 (5.64-13.70) |
| Day 3 | 10.42 (6.82-15.92) |
| Day 7 | 9.89 (6.80-14.66) |

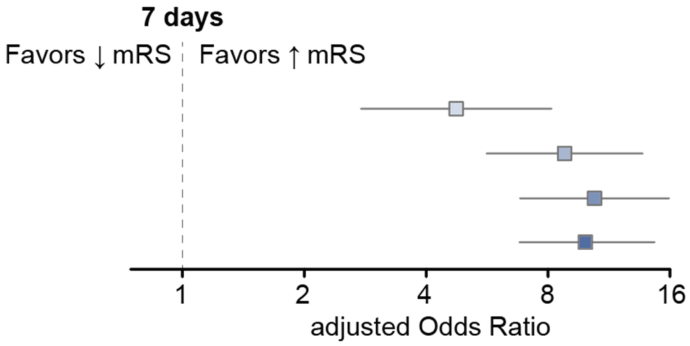

Shown is a forest plot of the associations of BD-tau levels upon admission, at day 2, day 3, and day 7 with mRS scores at 7 days. The odds ratios (95% confidence interval) were calculated using a multivariable logistic regression model adjusting for age, sex, hypertension, and the premorbid mRS score. mRS, modified Rankin Scale; aOR, adjusted Odds Ratio; CI, Confidence Interval; BD-tau, brain-derived tau.

**eFigure 15. Association of BD-tau over time after stroke with the rate of functional independence at 90 days.**

| BD-tau sampling | aOR (95% CI) |
|---|---|
| Upon admission | 5.84 (2.50-14.45) |
| Day 2 | 7.62 (3.96-15.42) |
| Day 3 | 9.69 (5.05-19.65) |
| Day 7 | 7.40 (4.21-13.61) |

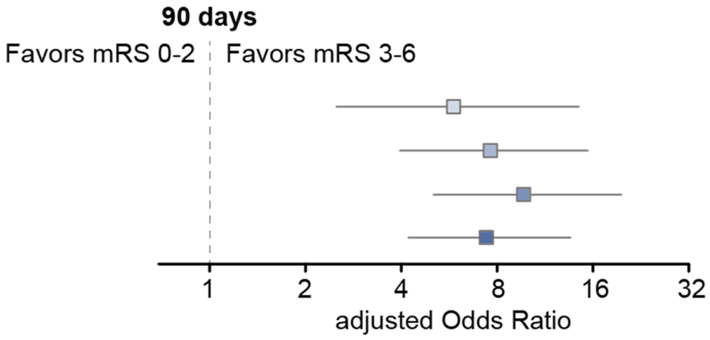

Shown is a forest plot of the associations of BD-tau levels upon admission, at day 2, day 3, and day 7 with the rate of functional dependence (mRS 3-6) vs. functional independence (mRS 0-2) at 90 days. The odds ratios (95% confidence interval) were calculated using a multivariable logistic regression model adjusting for age, sex, hypertension, and the premorbid mRS score. mRS, modified Rankin Scale; aOR, adjusted Odds Ratio; CI, Confidence Interval; BD-tau, brain-derived tau.

### eFigure 16. Association of BD-tau over time after stroke with the rate of functional independence at day 7.

| BD-tau sampling | aOR (95% CI) |
|---|---|
| Upon admission | 6.68 (3.23-14.42) |
| Day 2 | 9.87 (5.49-18.54) |
| Day 3 | 11.17 (6.34-20.60) |
| Day 7 | 10.57 (6.34-18.35) |

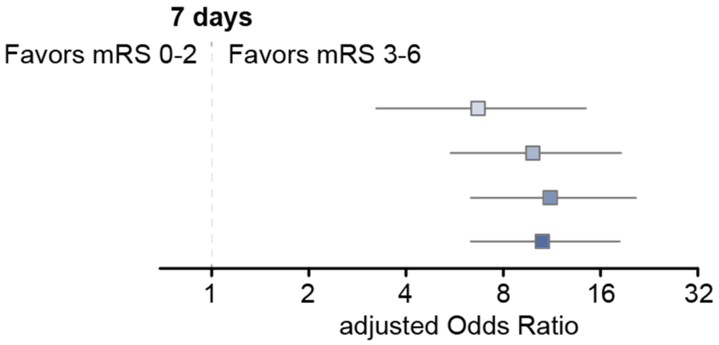

Shown is a forest plot of the associations of BD-tau levels upon admission, at day 2, day 3, and day 7 with the rate of functional dependence (mRS 3-6) vs. functional independence (mRS 0-2) at 90 days. The odds ratios (95% confidence interval) were calculated using a multivariable logistic regression model adjusting for age, sex, hypertension, and the premorbid mRS score. mRS, modified Rankin Scale; aOR, adjusted Odds Ratio; CI, Confidence Interval; BD-tau, brain-derived tau.

### eFigure 17. Variable importance for the 90-day mRS score in analysis restricted to patients with MRI-based infarct volumetry

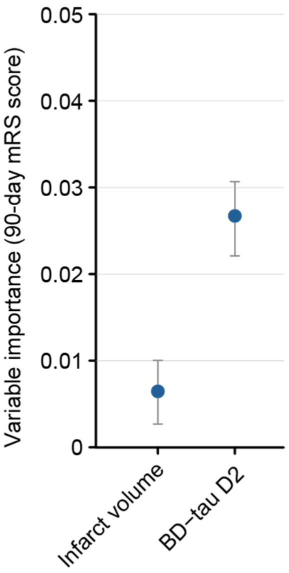

Shown are the median (dot) and 5% and 95% quantiles (whiskers) of importance values to predict the 90-day mRS score in a multivariable random forest regression model. BD-tau, brain-derived tau; mRS, modified Rankin Scale; MRI, magnetic resonance imaging.

### 3 eTables

### eTable 1. Concentrations of plasma BD-tau in quality controls

| | **Quality control 1 (n = 76)** | **Quality control 2 (n =78)** |
|---|---|---|
| Mean concentration (pg/ml) | 3.7 ±1.32 | 1.2 ±0.46 |
| Repeatability (%CV) | 8.5 | 9.1 |

BD-tau, brain-derived tau; CV, coefficient of variation.

### eTable 2. Discharge diagnoses of patients with stroke mimics

| Diagnosis | Patients, No. (%) |
|---|---|
| Epileptic seizure | 29 (57) |
| Focal neuropathy | 4 (8) |
| Electrolyte imbalance | 3 (6) |
| Neurodegenerative disease | 3 (6) |
| Septic encephalopathy | 3 (6) |
| Posterior reversible encephalopathy syndrome | 2 (4) |
| Somatization | 1 (2) |
| Syncope | 1 (2) |
| Peripheral vertigo | 1 (2) |
| Migraine | 1 (2) |
| Hyperglycemia | 1 (2) |
| Bacterial meningitis | 1 (2) |
| Brain neoplasm | 1 (2) |

### eTable 3. Representativeness of study participants

| Category | Considerations |
|---|---|
| Disease under investigation | Patients with acute ischemic stroke. |
| Sex and gender | Women present slightly less often with stroke than men (portion ranging from 43 % to 51 %).[11,12] |
| Age | Patients presenting with stroke have a median age of around 75 years.[11,12] |
| Race or ethnic group | Patients presenting with stroke in Germany are almost exclusively of Caucasian origin. |
| Pre-stroke status | Patients presenting with stroke have a high pre-stroke functional status (median pre-stroke modified Rankin Scale score 0-1), show a high prevalence of hypertension (~ 75%), diabetes mellitus (~21%), and atrial fibrillation (~24%).[12] |
| Stroke severity | Patients presenting with stroke have a median National Institutes of Health Stroke Scale score of 5-6.[11-13] |
| Stroke treatment | In a recent nationwide analysis from the United States, the rate of intravenous thrombolysis in patients presenting with stroke was 10%. The rate of endovascular treatment was 3%.[14] The rate of any acute reperfusion therapy was 40% in a recent nationwide analysis from Switzerland.[12] |
| Time to admission | The median time from stroke onset to hospital admission differs widely between 2 and 24 hours depending on the stroke population and setting studied.[15] The median time from stroke onset to hospital admission in the nationwide swiss stroke registry was approx. 4 hours.[12] |
| Overall representativeness of this study | The participants in the present study demonstrated a higher than expected ratio of men to women. The mean age, pre-stroke status, and stroke severity in the study population are consistent with prior studies from Europe and the United States. The time from stroke onset to admission was similar to other reports presenting procedural metrics of comprehensive stroke centers. Our study population received a slightly higher rate of acute reperfusion therapies than expected. |
